# Functional Assessment and Impact of Seizures on Cognitive Outcome in a Cohort of Individuals with KBG Syndrome

**DOI:** 10.1101/2024.04.17.24305757

**Authors:** Kathleen P Sarino, Lily Guo, Edward Yi, Jiyeon Park, Ola Kierzkowska, Drake Carter, Elaine Marchi, Gholson J Lyon

**Affiliations:** Department of Human Genetics, NYS Institute for Basic Research in Developmental Disabilities,1050 Forest Hill Road, Staten Island NY 10314, United States of America; George A. Jervis Clinic, NYS Institute for Basic Research in Developmental Disabilities, 1050 Forest Hill Road, Staten Island NY 10314, United States of America; Biology PhD Program, The Graduate Center, The City University of New York, New York, United States of America

**Keywords:** *ANKRD11* gene, epilepsy, KBG syndrome, neurodevelopment, seizures

## Abstract

**OBJECTIVE:** This study aimed to further examine the impact of epileptic seizures on neurocognitive outcomes in KBG syndrome, a rare genetic neurodevelopmental disorder characterized by pathogenic variants in the gene ANKRD11.

**METHODS:** A single clinician interviewed a cohort of individuals with genetically confirmed cases of KBG syndrome. Medical records and other relevant data were collected for each participant. To evaluate participants’ adaptive functioning, trained professionals conducted assessments using the Vineland–3 Adaptive Behavior Scales. The assessment compared individuals with epilepsy to those without seizures and covered the domains of communication, daily living skills, socialization, and maladaptive behaviors. Further comparisons were drawn based on insights from interviews and information extracted from participants’ medical records.

**RESULTS:** Thirty-nine individuals (22 males, 17 females) with KBG syndrome, confirmed through genetic analysis, were interviewed via videoconferencing by a single physician, followed by Vineland-3 assessment by trained raters. Individuals with KBG syndrome came from 36 unique families spanning 11 countries. While the KBG cohort displayed lower overall adaptive behavior composite scores compared to the average population, several members displayed standard scores at or higher than average, as well as higher scores compared to those with the neurodevelopmental disorder Ogden syndrome. Within the KBG cohort, males consistently scored lower than females across all domains, but none of these categories reached statistical significance. While the group with epilepsy exhibited overall lower scores than the non-seizure group in every category, statistical significance was only reached in the written communication subdomain. We predict this lack of significance is limited by low sample size, reducing study power.

**CONCLUSIONS:** Due to the rarity of KBG syndrome, our research provides valuable insights that can aid in epilepsy screening and inform assessment strategies for neurocognitive functioning in those with this condition. The cohort performed overall higher than expected with outliers existing in both directions. Although our results suggest that seizures might influence the trajectory of KBG syndrome, the approaching but overall absence of statistical significance between study groups underscores the necessity for a more extensive cohort to discern subtle variations in functioning. Conducting Vineland–3 assessments in the KBG syndrome population can enhance research insights regarding differences between those with and without epilepsy. Given the data collected, we recommend vigilant monitoring for seizures following a KBG diagnosis, with consideration for performing baseline EEG assessments.

## Introduction

KBG syndrome is a neurodevelopmental disorder characterized by mutations in the ankyrin repeat domain 11 gene (*ANKRD11*) or microdeletions that affect *ANKRD11* on chromosome 16q24.3.^1–6^ While the precise functional role of ANKRD11 in the brain remains unclear, variants in this gene have been associated with cognitive and neurological anomalies. These include a range of seizures such as focal and generalized seizures, tonic-clonic and absence seizures, myoclonic seizures, and unclassified sleep-related seizures.^7^ Some individuals may exhibit electroencephalogram (EEG) abnormalities without clinically evident seizures.^8^ Those with KBG syndrome are more susceptible to epilepsy, which occurs in a significant percentage of cases,^9^ although epilepsy is not formally recognized as a defining characteristic of this condition.

It is well-established in various epileptic encephalopathies that the onset of seizures can trigger developmental regression or be linked to a poorer prognosis.^10^ Similarly, other genetic neurodevelopmental conditions such as Rett syndrome display a high incidence of epilepsy associated with more severe developmental disabilities.^11^ The impact of seizures on cognitive outcomes has been a significant focus of research.^12,13^ Often, individuals with early-onset or prolonged epilepsy experience cognitive impairments, including disruptions in memory, attention, processing speed, and executive functioning.^14,15^ The type and location of seizures can also influence the extent of cognitive impairment. Previous studies have hypothesized a potential link between early onset of seizures and a more severe form of KBG syndrome.^16^

In this study, we evaluate cognitive outcomes in individuals with KBG syndrome, both with and without seizures, using the Vineland Adaptive Behavior Scales, Third Edition (Vineland–3).^17,18^ This tool is a standardized formal assessment used to measure adaptive behavior in the diagnosis of intellectual and developmental disabilities or delays. We aim to explore potential phenotype correlations through analysis of communication, socialization, and daily living skills, focusing on how these aspects may be influenced by the presence of epilepsy in KBG syndrome.

## Methods

### Participants

Thirty-nine individuals (22 males, 17 females) with KBG syndrome, confirmed through genetic analysis, were interviewed via videoconferencing by a single physician board certified in child and adolescent psychiatry (GJL). These individuals belonged to 36 unique families spanning 11 countries. Prior to the initial interviews, medical records including genetic reports, facial, and whole-body photographs were collected from participants. Videoconferencing occurred in two phases: preliminary data collection (GJL) and Vineland–3 adaptive behavior assessment (KS,LG,EY,JP). All sessions spanned from one to four hours and were conducted in English, with a translator used for one family whose primary language was Spanish. With prior consent, interviews were recorded and archived for subsequent analyses.

Twenty-two individuals in our cohort have not been previously published in the literature (KBG- GJL-001 to KBG-GJL-005, KBG-GJL-007 to KBG-GJL-013, KBG-GJL-016, KBG-GJL-018 to KBG-GJL-020, KBG-GJL-023 to KBG-GJL-025, and KBG-GJL-029, KBG-GJL-039, and KBG-GJL-040). Four affected individuals (KBG-GJL-007, KBG-GJL-010, KBG-GJL-032, and KBG-GJL-042) had one or more children who also possessed a confirmed KBG diagnosis. At least one child of each of these four participants was part of the assessed cohort. These identifiers (such as KBG-GJL-040) are not known to anyone outside of the study team, thus decreasing risk of re-identification.

### Data Collection

*Phase 1.* Initial videoconferences were held over 20 months from February 2021 to October 2022. Families were recruited through a KBG syndrome Facebook group or referrals of other families with known members possessing KBG syndrome. ANKRD11 variants were annotated to the NM_013275.5 transcript in GrCh37/hg19. The severity of developmental delay and level of functioning was assessed by the physician through patient interaction, paternal/maternal reports, and cross-referencing existing medical records. Metrics were systematically documented, encompassing speech and motor delays, behavioral issues, and neurological abnormalities, including seizures. The type and onset of epilepsy, along with comprehensive details about the treatment, such as the specific medications used, were recorded. Medical records pertaining to epileptic events in relevant individuals were thoroughly reviewed. Any abnormalities detected through neuroimaging, such as MRI and EEG, were noted, and families were requested to provide copies of these records. Furthermore, additional neurological features, such as abnormal pain thresholds and altered tactile sensations, were also documented, as well as other features such as anatomical abnormalities or complications during birth.

*Phase 2.* Adaptive behavior was assessed using the Vineland–3, administered through teleconferences from September 2022 to January 2023 using a small group of trained raters (KS,LG,EY,JP). Each assessment involved a semi-structured interview with a parent or primary caregiver knowledgeable about the daily behaviors of the individual with KBG syndrome. The scores obtained from the Vineland–3 for those with KBG syndrome were compared to those of the general population and individuals with another genetic neurodevelopmental disorder, Ogden syndrome, characterized by pathogenic variants in the NAA10 and NAA15 genes.^19,20^ Vineland– 3 scores of those with Ogden Syndrome were collected previously through videoconference in a similar manner as the KBG syndrome data collection.

There was one family, possessing identifier KBG-Family-24 and consisting of a mother and her two sons, also previously published^16^, who was initially included in all analyses, but was then later removed (with all analyses performed again) due to having a variant of uncertain significance (VUS) (consisting of a Val586Met missense mutation). Exome re-analysis of this trio is underway, in search for any other possible cause of the condition in that family.

### Data Analysis

The Vineland–3 evaluates adaptive behavior in three major domains: communication, daily living skills, and socialization **(Figure 1),** each with a normative mean of 100 and a standard deviation of 15. These domains are further divided into subdomains. In addition to the three main domains, the Vineland provides an option for additional assessment through the maladaptive behavior domain, which focuses on internalizing behaviors such as anxiety and depression as well as externalizing behaviors such as hyperactivity and disruptive behavior. Sub-scores from each domain contribute to the overall Adaptive Behavior Composite (ABC) score, reflecting the individual’s overall comprehensive adaptive functioning.

**Figure 1:**
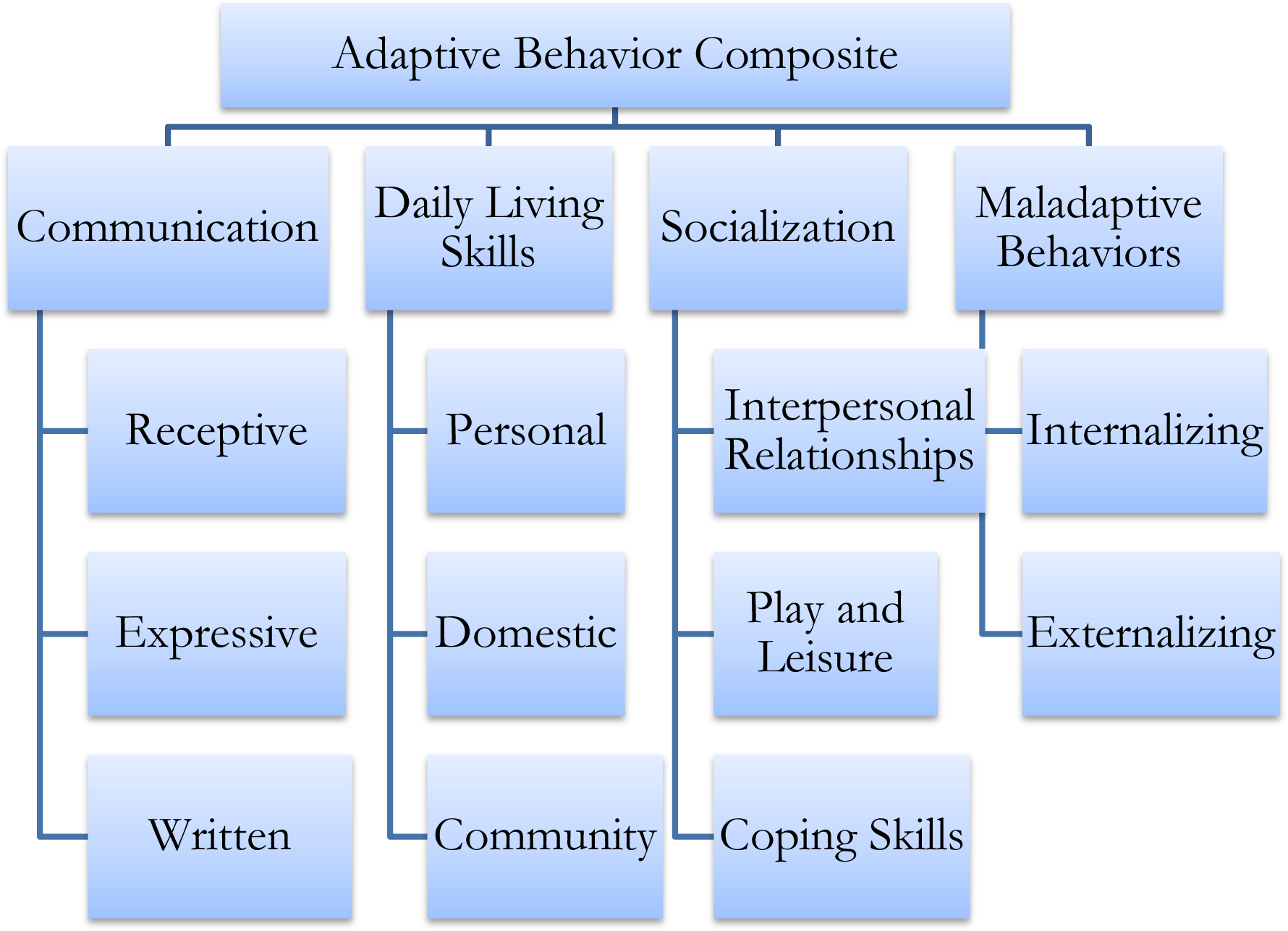
Vineland–3 Format: Domains and Subdomains. The Vineland–3 consists of three major domains, each containing three subdomains. The additional maladaptive behavior section includes two subdomains, internalizing and externalizing.

Vineland–3 data was analyzed using Prism GraphPad (version 9.5.1). Descriptive statistics, including mean, standard deviation, and range were computed for each of the three core domains and the ABC score. Independent two tailed t-tests were used to compare differences across domain and ABC scores. Separate analyses for sex were completed with an F-test. Simple linear regression was used to assess the effects of age on ABC score. Comparative analysis was conducted using one-sample t-tests for epilepsy versus no seizures. Effect sizes were calculated to determine the practical significance of any observed differences.

### Results

The demographics of the cohort are displayed in **Table 1**. The locations of ANKRD11 variants are depicted in **Figure 2** with specific mutations described in **Table 2**. 17 individuals had de novo variants while 17 had unknown modes of inheritance. Mutations in three individuals were maternally inherited while two were paternally inherited. When analyzing types of mutations, 23 possessed frameshift mutations, 11 had nonsense mutations, one had a missense mutation, and the remaining four had copy number variants involving deletions of the gene. Mutations were classified per the American College of Medical Genetics and Genomics (ACMG) criteria for the interpretation of sequence variants.^21^ All participants analyzed were either class one pathogenic (n = 36) or class two likely pathogenic (n = 3).

**Fig 2.**
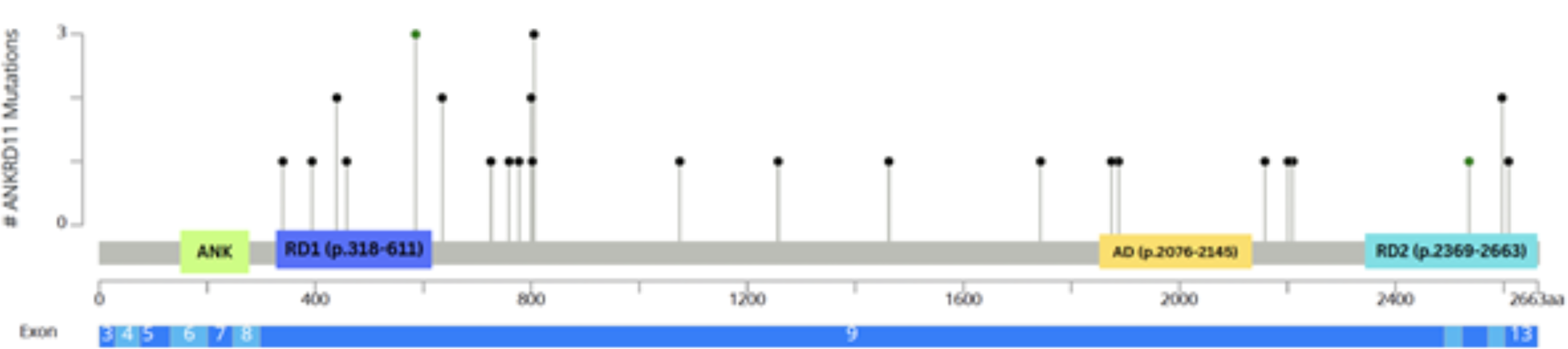
KBG syndrome cohort mutations. The coding exons for ANKRDII are depicted to scale. Abbreviations: aa amino acid. The figure was made using: https://www.cbioPortal.org/mutation_mapper.

**Table 1:** Demographics.

| **Table 1: Demographics** |  |  |
| --- | --- | --- |
| **Factor** | **Level** | **N** |
| Total |  | 39 |
| Sex | Male | 22 (56.4%) |
|  | Female | 17 (43.6%) |
| Mean Age (range) |  | 14.7 years (1, 61) |
| Race | Caucasian | 39 (100%) |
| Ethnicity | Non-Hispanic | 29 (74.4%) |
|  | Hispanic | 10 (25.6%) |
| Country of Origin | United States | 21 (53.8%) |
|  | Spain | 5 (12.8%) |
|  | United Kingdom | 4 (10.2%) |
|  | Germany | 2 (5.1%) |
|  | Australia | 1 (2.6%) |
|  | Canada | 1 (2.6%) |
|  | Mexico | 1 (2.6%) |
|  | Portugal | 1 (2.6%) |
|  | Argentina | 1 (2.6%) |
|  | Ecuador | 1 (2.6%) |
|  | Lebanon | 1 (2.6%) |

**Table 2:** Cohort Mutations and Classifications.

| Individual Identifier | Family Identifier | Age at time of video conference | Gender | g.DNA (Hg19/GRCh37) | cDNA (NM_013275.5) | Protein Effect | Variant type | Mode of Inheritance | Classification | ACMG/AMP Guidelines |
| --- | --- | --- | --- | --- | --- | --- | --- | --- | --- | --- |
| **KBG-GJL-001** | KBG-Family-1 | 0-4 years | Male | g.chr16:89351043-89351047 | c.1903_1907del | p.(Lys635fs) | frameshift | de novo | Class 1: pathogenic | PVS1 Very Strong; PS2 Strong; PM2 Moderate |
| **KBG-GJL-002** | KBG-Family-2 | 0-4 years | Female | g.16q24.3 | c.1181delA | p.(Asn394Ilefs*33) | frameshift | de novo | Class 1: pathogenic | PVS1 Very Strong; PS2 Strong; PM2 Moderate |
| **KBG-GJL-003** | KBG-Family-3 | 11-15 years | Male | g.Chr16:89346478 | c.6472G>T | p.(Glu2158*) | nonsense | de novo | Class 1: pathogenic | PVS1 Very Strong; PS2 Strong; PM2 Moderate |
| **KBG-GJL-004** | KBG-Family-4 | 21-25 years | Male | g.unknown | c.5620dupA | p.Thr1874fs | frameshift | unknown | Class 1: pathogenic | PVS1 Very Strong; PM2 Moderate |
| **KBG-GJL-005** | KBG-Family-5 | 5-10 years | Female | g.unknown | 334 kb loss in 16q24.3 | - | copy number variant | de novo | Class 1: pathogenic | PVS1 Very Strong; PP2 supporting |
| KBG-GJL-006 | KBG-Family-6 | 5-10 years | Female | NC_000016.9:g.89350544_89350547del | c.2404_2407del | p.(Leu802Lysfs*60) | frameshift | de novo | Class 1: pathogenic | PVS1 Very Strong; PS2 Strong; PM2 Moderate |
| **KBG-GJL-007** | KBG-Family-7 | 26-30 years | Female | g.Chr 16:89495519_89542025 | 34Kb loss in 16q24.3 | - | copy number variant | unknown | Class 1: pathogenic | PVS1 Very Strong; PP1 supporting; PP2 supporting |
| **KBG-GJL-008** | KBG-Family-7 | 0-4 years | Male | g.89495519_89542025 | 46.5Kb loss in 16q24.3 | - | copy number variant | maternal | Class 1: pathogenic | PVS1 Very Strong; PP1 supporting, PP2 supporting |
| **KBG-GJL-009** | KBG-Family-8 | 5-10 years | Male | g.unknown | 158 kB loss in 16q24.3 | - | copy number variant | unknown | Class 1: pathogenic | PVS1 Very Strong; PP2 supporting |
| **KBG-GJL-010** | KBG-Family-9 | 26-30 years | Female | g.Chr16-89350522 | c.2413_2416 | p.(Glu0805fs) | frameshift | unknown | Class 1: pathogenic | PVS1 Very Strong; PM2 Moderate |
| **KBG-GJL-011** | KBG-Family-9 | 0-4 years | Male | g.Chr16-89350522 | c.2413_2416 | p.(Glu0805fs) | frameshift | maternal | Class 1: pathogenic | PVS1 Very Strong; PS2 Strong; PM2 Moderate |
| **KBG-GJL-012** | KBG-Family-10 | 5-10 years | Male | g.89346322 C>A | c.6628G>T | p.(Glu2210*) | nonsense | unknown | Class 1: pathogenic | PVS1 Very Strong; PM2 Moderate |
| **KBG-GJL-013** | KBG-Family-11 | 5-10 years | Male | g.unknown | c.2398_2401delG AAA | p.(Glu800Asn fs*62) | frameshift | de novo | Class 1: pathogenic | PVS1 Very Strong; PS2 Strong; PM2 Moderate |
| KBG-GJL-014 | KBG-Family-12 | 21-25 years | Female | NC_000016.9:g.89350773_89350774del | c.2177_2178del | p.(Lys0726Arg fs*15) | frameshift | de novo | Class 1: pathogenic | PVS1 Very Strong; PS2 Strong; PM2 Moderate |
| KBG-GJL-015 | KBG-Family-13 | 21-25years | Male | g.unknown | c.1018dup | p.(Thr340Asn fs*9) | frameshift | unknown | Class 1: pathogenic | PVS1 Very Strong; PM2 Moderate |
| **KBG-GJL-016** | KBG-Family-14 | 5-10 years | Female | g.Chr16:89351042 | c.1903_1907delA AACA | p.(Lys0635Gln fs*26) | frameshift | unknown | Class 1: pathogenic | PVS1 Very Strong; PM2 Moderate |
| KBG-GJL-017 | KBG-Family-15 | 11-15 years | Male | NC_000016.9:g.89335053G>A | c.7825C>T | p.(Gln2609*) | nonsense | de novo | Class 1: pathogenic | PVS1 Very Strong; PS2 Strong; PM2 Moderate |
| **KBG-GJL-018** | KBG-Family-16 | 5-10 years | Male | g.unknown | c.1318C>T | p.(Arg0440*) | nonsense | unknown | Class 1: pathogenic | PVS1 Very Strong; PM2 Moderate |
| **KBG-GJL-019** | KBG-Family-17 | 0-4 years | Female | g.16:89351632G>A | c.1318C>T | p.(Arg440*) | nonsense | de novo | Class 1: pathogenic | PVS1 Very Strong; PS2 Strong; PM2 Moderate |
| **KBG-GJL-020** | KBG-Family-18 | 5-10 years | Female | g.89350540_89350543delTTTT | c.2409_2412 del | p.(Glu0805Argfs*57) | frameshift | unknown | Class 1: pathogenic | PVS1 Very Strong; PM2 Moderate |
| KBG-GJL-021 | KBG-Family-19 | 5-10 years | Male | NC_000016.9:g.89349180_89349181del | c.3770_3771del | p.(Lys1257Argfs*25) | frameshift | de novo | Class 1: pathogenic | PVS1 Very Strong; PS2 Strong; PM2 Moderate |
| KBG-GJL-022 | KBG-Family-20 | 5-10 years | Male | NC_000016.9:g.89341328C>T | c.7607G>A | p.(Arg2536Gln) | missense | de novo | Class 2: Likely pathogenic | PS2 Strong, PM2 Moderate |
| **KBG-GJL-023** | KBG-Family-21 | 21-25 years | Male | Chr16:89282157 | c.4384dupA | p.(Arg1462Lysfs*92) | frameshift | unknown | Class 1: pathogenic | PVS1 Very Strong; PM2 Moderate |
| **KBG-GJL-024** | KBG-Family-22 | 5-10 years | Female | g. Chr16:89.284.140-89.284.144 | c.2398_2401delGAAA | p.(Glu800Asnfs*62) | frameshift | unknown | Class 1: Pathogenic | PVS1 Very Strong; PM2 Moderate |
| **KBG-GJL-025** | KBG-Family-23 | 0-4 years | Male | g.Chr16 | c.1372C>T | p.(Arg458*) | nonsense | paternal | Class 1: Pathogenic | PVS1 Very Strong; PM2 Moderate |
| **KBG-GJL-029** | KBG-Family-25 | 6-10 years | Female | g.unknown | c.2329_2332del | p.(Glu777Argfs*5) | frameshift | unknown | Class 1: pathogenic | PVS1 Very Strong; PM2 Moderate |
| KBG-GJL-030 | KBG-Family-26 | 0-4 years | Male | NC_000016.9:g.89350777_89350780del | c.2175_2178del | p.(Asn0725Lysfs*23) | frameshift | de novo | Class 1: pathogenic | PVS1 Very Strong; PS2 Strong; PM2 Moderate |
| KBG-GJL-031 | KBG-Family-27 | 21-25 years | Male | NC_000016.9:g.89337242T>A | c.7789A>T | p.(Lys2597*) | nonsense | paternal | Class 2: Likely pathogenic | PVS1 Very Strong; PM2 Moderate |
| KBG-GJL-032 | KBG-Family-27 | 55-60 years | Male | NC_000016.9:g.89337242T>A | c.7789A>T | p.(Lys2597*) | nonsense | unknown | Class 2: Likely pathogenic | PVS1 Very Strong; PM2 Moderate |
| KBG-GJL-033 | KBG-Family-28 | 11-15 years | Female | NC_000016.9:g.89350677dup | c.2273dup | p.(Arg759Glu*23fs) | frameshift | unknown | Class 1: pathogenic | PVS1 Very Strong; PM2 Moderate |
| KBG-GJL-034 | KBG-Family-29 | 21-25 years | Female | NC_000016.9:g.89346353_89346354insT | c.6596_6597insA | p.(Ala2201Cysfs*6) | frameshift | de novo | Class 1: pathogenic | PVS1 Very Strong; PS2 Strong; PM2 Moderate |
| KBG-GJL-035 | KBG-Family-30 | 11-15 years | Female | NC_000016.9:g.89347291G>A | c.5659C>T | p.(Gln1887*) | nonsense | unknown | Class 1: pathogenic | PVS1 Very Strong; PM2 Moderate |
| KBG-GJL-036 | KBG-Family-31 | 5-10 years | Male | NC_000016.9:g.89347723G>A | c.5227C>T | p.(Gln1743*) | nonsense | maternal | Class 1: pathogenic | PVS1 Very Strong; PM2 Moderate |
| KBG-GJL-037 | KBG-Family-32 | 16-20 years | Male | NC_000016.9:g.89349727_89349730del | c.3224_3227del | p.(Glu1075Glyfs*242) | frameshift | unknown | Class 1: pathogenic | PVS1 Very Strong; PM2 Moderate |
| KBG-GJL-038 | KBG-Family-33 | 5-10 years | Male | NC_000016.9:g.89350540_89350543del | c.2409_2412del | p.(Glu805Argfs*57) | frameshift | de novo | Class 1: pathogenic | PVS1 Very Strong; PS2 Strong; PM2 Moderate |
| **KBG-GJL-039** | KBG-Family-34 | 16-20 years | Male | g.Chr16 | c.3770-3771del | p.Lys1257Argfs*25 | frameshift | de novo | Class 1: pathogenic | PVS1 Very Strong; PS2 Strong; PM2 Moderate |
| **KBG-GJL-040** | KBG-Family-35 | 0-4 years | Male | g.16:8934853 | c.4396_4397 | p.Arg1466Glyfs*87 | frameshift | de novo | Class 1: pathogenic | PVS1 Very Strong; PS2 Strong; PM2 Moderate |
| KBG-GJL-041 | KBG-Family-36 | 16-20 years | Female | NC_000016.9:g.89351062del | c.1893del | p.(Lys0631fs) | frameshift | de novo | Class 1: pathogenic | PVS1 Very Strong; PS2 Strong; PM2 Moderate |
| KBG-GJL-042 | KBG-Family-31 | 30-35 years | Female | NC_000016.9:g.89347723G>A | c.5227C>T | p.(Gln1743*) | nonsense | unknown | Class 1: pathogenic | PVS1 Very Strong; PM2 Moderate |

Participant mutations and classifications based on ACMG guidelines. Participants in bold have previously been published in a paper by Guo et al (1). One family, possessing identifier KBG-Family-24 and consisting of a mother and her two sons, was removed from all data analyses due to having a variant of uncertain significance (VUS).

### Cohort Analysis

While the cohort displayed a lower mean in ABC scores compared to the average population standard mean of 100, several members displayed scores at or higher than average, as well as higher scores compared to those with Ogden syndrome and closer to those with NAA15- related neurodevelopmental syndrome^20,22,23^ (**Figure 3A)**. While KBG syndrome females had a higher mean ABC score when compared to males, analyses by t-test revealed no statistically significant difference (**Figure 3B**). Additional comparisons by sex were made including ABC scores based on age (**Figure 3C and 3D**). This graphical representation of ABC scores showed consistent values with a marginally positive trend, presenting a slightly negative slope for males versus a slightly positive slope for females. This stratification by both sex and age did not reach statistical significance when compared (**Figure 3C and 3D**).

**Figure 3:**
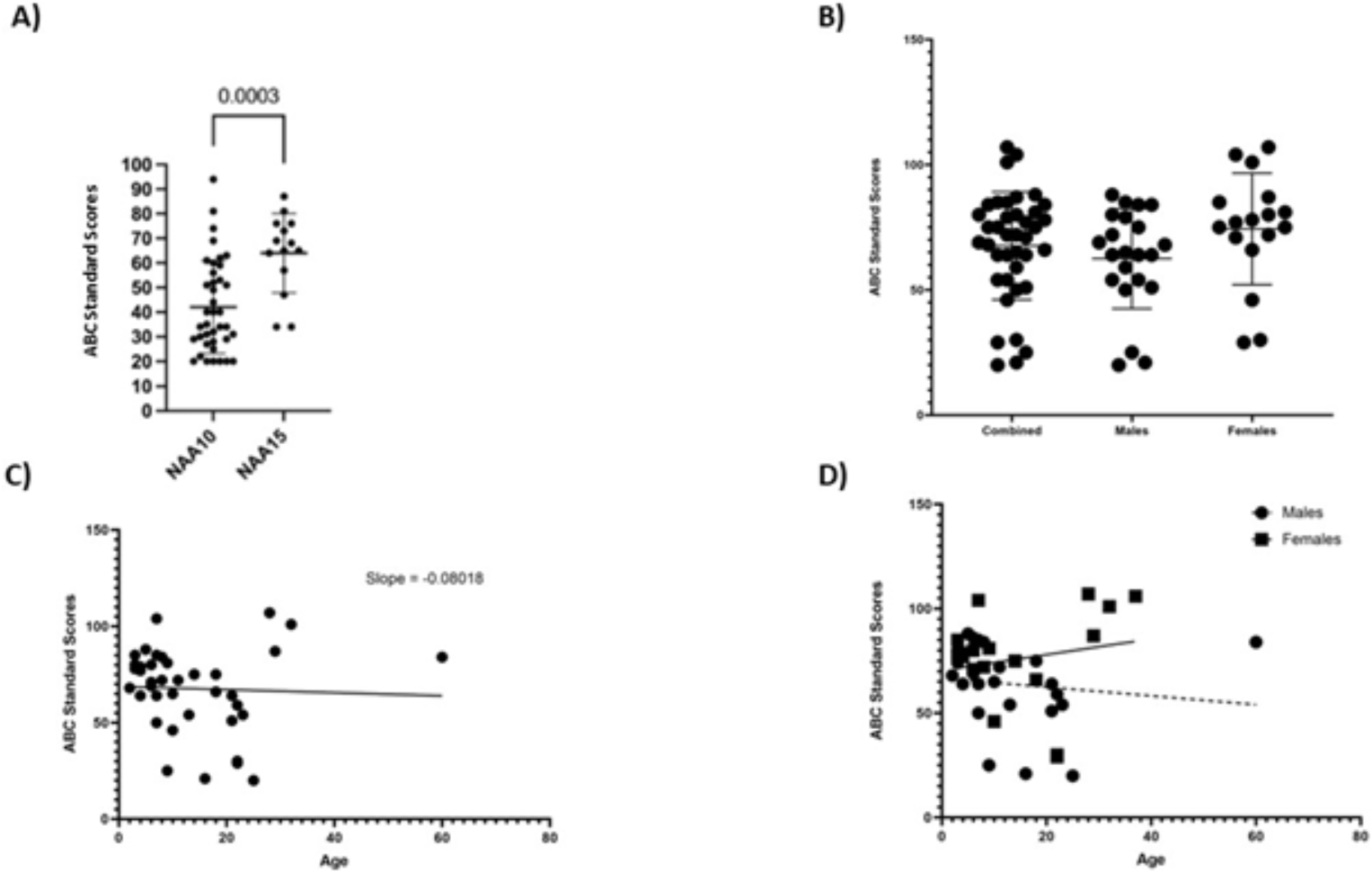
Vineland Results in KBG ts. Other Neurodevelopmental Conditions. A) ABC Standard scores for Ogden (NAA15 variants) and NAA15-related neurodevelopmental syndrom e B) ABC scores for KBG cohort (mean score αf 67.67) including separate m eans for males (62.50) and fern ales (7435), p= 0.0887 C) ABC scores for KBG cohort overtime D) Comparison of ABC scores over time between males (slope = -0.1370 andfemales (slope = 0.0405), notfoundtohave a statistically significant difference (p = 0 3 3 43).

Mean domain scores were similar to ABC scores, averaging around a score of 70 (**Figure 4**). Specific domain scores are shown in **Table 3**. Within the cohort, males consistently scored lower than females across all categories, but none of the domains reached statistical significance (**Table 3**).

**Figure 4:**
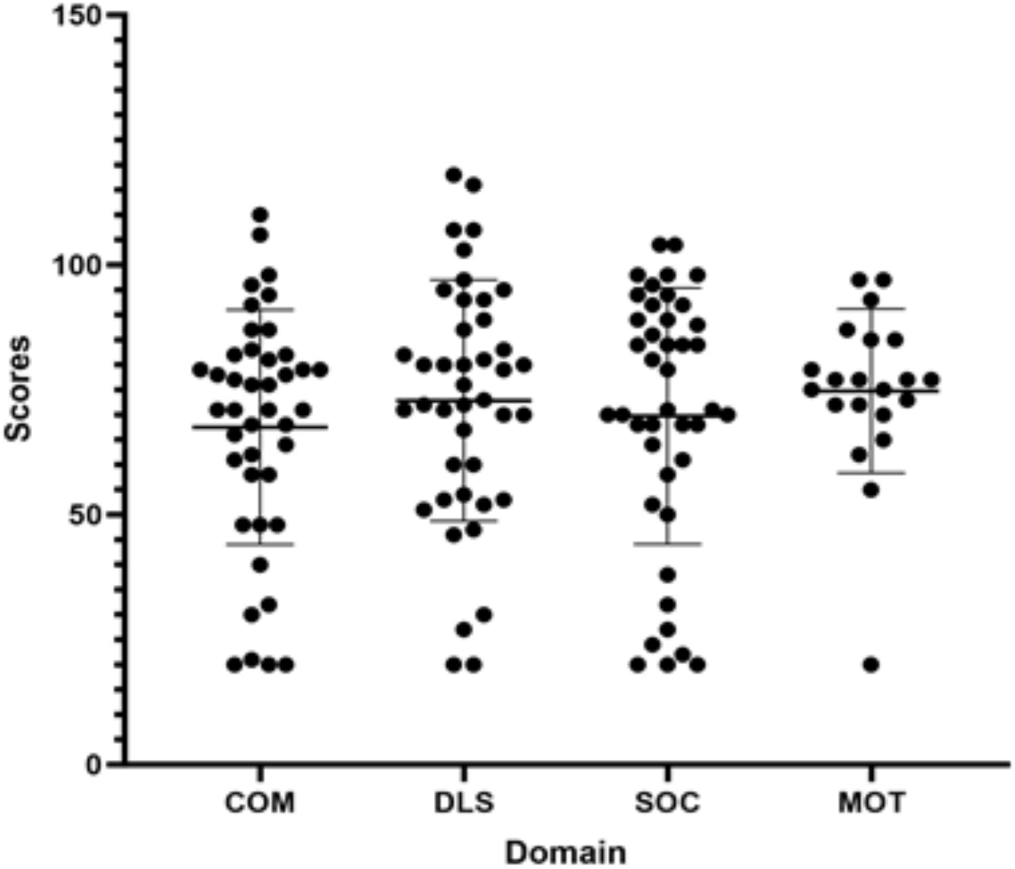
Domain Scores of full cohort. Core domain scores including means for total cohort.

**Table 3:** Domain Scores by Sex.

| Table 3: Domain Scores by Sex |  |  |  |  |
| --- | --- | --- | --- | --- |
|  | COM (SD) | DLS (SD) | SOC (SD) | MOT (SD) |
| **Total** | 65.51 (22.91) | 71.95 (24.37) | 67.95 (25.64) | 74.47 (17.12) |
| **Male** | 59.64 (21.79) | 65.41 (22.89) | 64.00 (25.99) | 71.36 (21.52) |
| **Female** | 73.12 (22.68) | 80.41 (24.23) | 73.06 (25.02) | 78.75 (7.498) |
| **p** | 0.0677 | 0.0553 | 0.2798 | 0.3682 |
| Overall mean domain scores and scores by male versus female.<br>(COM: communication; DLS: daily living skills; SOC: social skills; MOT: motor skills) |  |  |  |  |

### Epilepsy Analysis

Participants with epilepsy are detailed in **Table 4**. Most epileptic patients experienced generalized tonic-clonic seizures, with absence seizures being the second most prevalent. When comparing the age of onset of seizures and whether initial age had any influence on adaptive scores, no statistically significant differences were indicated (**Figure 5 and Table 5**). Six interviewed individuals in our cohort reported one or more possible or potential seizures that did not clearly exhibit epilepsy descriptions and were not diagnosed by a clinician. This includes participant KBG-GJL-014 whose mother reported occasional “zoning out” for short periods, possibly suggesting absence seizures, but was not certain or confirmed. KBG-GJL-007 mentioned a memory of a possible seizure in childhood but was also unsure of its nature. All six individuals who experienced a single, unconfirmed seizure or had suspected but undiagnosed seizures did not fulfill epilepsy criteria and were excluded from the following seizure analysis. They were not included in either the epilepsy group (n = 11) or the "non-seizure” group (n = 28) given the questionable history.

**Figure 5:**
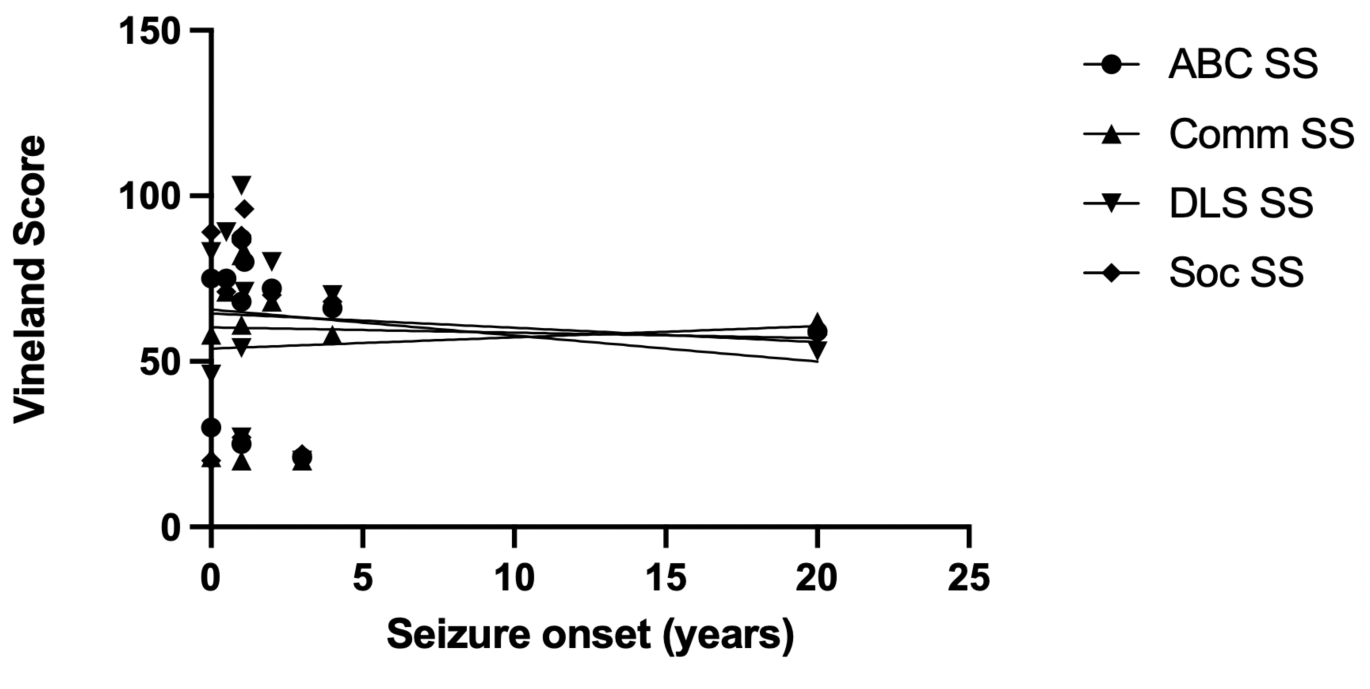
V**i**neland**–3 Score vs. Seizure Onset.** Comparison of ABC standard scores and domain scores for participants compared to seizure onset. No statistical significance was achieved.

**Table 4:** Seizure Analysis.

| **Table 4: Seizure Analysis** |  |  |  |  |  |
| --- | --- | --- | --- | --- | --- |
| **Identifier** | **Onset** | **Type(s) of Seizures** | **Treatment** | **Neuroimaging** | **Subjective Comments Per Primary Caretaker** |
| **KBG-GJL-003** | 0-5 years old | Absence; tonic- clonic with loss of bladder control | Levetiracetam, lamotrigine*, topiramate*, sodium valproate*, clobazam* | EEG shows frequent frontally per dominant generalized spike, slow wave discharges; Slowing of EEG background and mild diffuse slowing; Findings consistent with symptomatic epilepsy, most likely tonic seizures | Felt seizures have affected development, with regression associated with seizure activity |
| **KBG-GJL-004** | 20-25 years old | Tonic-clonic | Levetiracetam* | EEG and MRI brain unremarkable | - |
| **KBG-GJL-006** | 0-5 years old | Myoclonic; complex partial; verbal tonic seizures with respiratory distress | Lacosamide*, rufinamide*, vagus nerve stimulator implanted at 0-5 years | Abnormal EEG<br>MRI brain shows arachnoid cyst | Felt onset of seizures halted normal development and resulted in regression |
| **KBG-GJL-010** | 0-5 years old | Generalized tonic-clonic | Carbamazepine | Abnormal EEG; MRI brain unremarkable | Felt seizures did not affect development |
| **KBG-GJL-011** | 0-5 years old | Generalized tonic-clonic | Levetiracetam, sodium valproate, topiramate | Normal EEG after first seizure; pathological EEG at 0-4 years<br>Brain MRI normal | No obvious change in trajectory or regression noted |
| **KBG-GJL-034** | 0-5 years old | Absence; generalized tonic-clonic | Lamotrigine* | Abnormal EEG | - |
| **KBG-GJL-035** | 0-5 years old | Myoclonic; generalized tonic-clonic | Sodium valproate, Lamotrigine* | EEG shows frequent generalized spike wave discharges; MRI brain shows multiple non-enhancing T2/FLAIR hyperintense subcortical lesions in left frontal lobe | - |
| **KBG-GJL-036** | 0-5 years old | Absence; generalized tonic-clonic | Sodium valproate* | EEG and MRI brain unremarkable | Felt seizures linked to increase in aggressive behaviors |
| **KBG-GJL-037** | 0-5 years old | Unspecified | Sodium valproate* | MRI brain shows light asymmetry in size of the temporal horn of left lateral ventricle (likely developmental), small pineal cyst | - |
| **KBG-GJL-038** | 0-5 years old | Generalized tonic-clonic, myoclonic, atonic, tonic, focal, absence | Clobazam*, lacosamide*, rufinamide* | EEG and MRI brain unremarkable | Felt massive regression was likely due to seizures |
| **KBG-GJL-041** | 0-5 years old | Generalized tonic-clonic | Lamotrigine* | MRI brain shows lateral right temporal bone lesion on squamosal petrosal suture<br>EEG (2010): Epileptiform discharges maximal over frontal regions but increased in sleep<br>EEG (2013): Normal sleep architecture with occasional epileptiform discharges | Unclear if seizures affected trajectory |

Participants in KBG cohort experiencing epileptic seizures (n = 11). Participants listed in bold have been previously published as part of the cohort in Guo et al (1).
\*: Patient's current medications taken at time of videoconference.

**Table 5:** Vineland–3 Standard Scores in Epilepsy Group.

| Table 5: Vineland-3 Standard Scores in Epilepsy Group. |  |  |  |  |
| --- | --- | --- | --- | --- |
|  | R2 | F statistic | Degree of freedom | P-value |
| ABC SS | 0.001585 | 0.01428 | 1, 9 | 0.9075 |
| Comm SS | 0.006783 | 0.06146 | 1, 9 | 0.8098 |
| DLS SS | 0.03039 | 0.2821 | 1, 9 | 0.6082 |
| Soc SS | 0.007818 | 0.07091 | 1, 9 | 0.7960 |
| ABC standard score, core domain scores, and subdomain scores were assessed for the epilepsy group (n = 11) and the non-seizure group (n = 28). |  |  |  |  |

Anecdotally, per primary caregivers, several participants (4 of the 11) indicated a level of developmental regression following onset of seizures. However, when comparing means of ABC standard scores, no statistical significance was found between those with and without epilepsy (**Figure 6**). Analysis of mean scores in the Vineland domains also revealed no statistical signficance **(Figure 7)**. Each of the domains were further assessed by subdomain, with the subdomain of written communication showing a statistically significant difference between epilepsy groups. The scores for communication subdomains are presented in **Figure 8**. The remaining subdomains (in the domains of daily living skills and socialization) as well as maladaptive behavior scores did not exhibit statistical significance between groups **(Figure 9)**.

**Figure 6:**
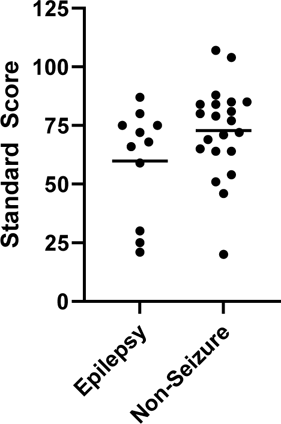
A**B**C **Standard Score of Epilepsy vs. Non-Seizure Groups.** The ABC score encompasses an individual’s overall adaptive functioning; the standard mean for a person without intellectual or developmental disabilities is 100 with a standard deviation of 15. The mean ABC standard score in the KBG epilepsy group is 59.82 and the mean of the non-seizure group is 72.86 with a p-value 0.105. * p < 0.05

**Figure 7:**
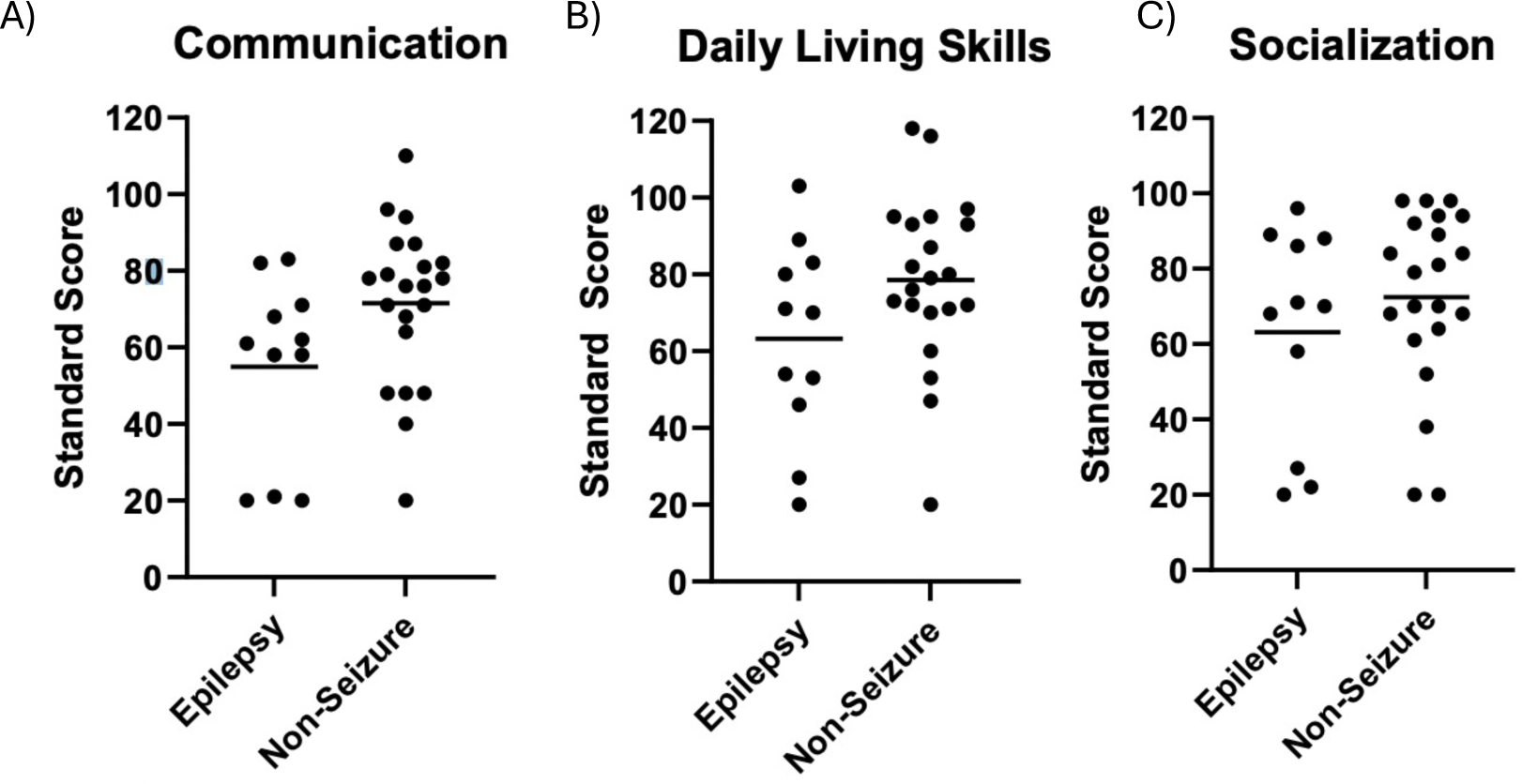
D**o**main **Scores of Epilepsy vs. Non-Seizure Groups.** The three major domains; the standard mean for a person without intellectual or developmental disabilities is 100 with a standard deviation of 15. **A** Mean communication domain standard score were 54.91 for the epilepsy group and 69.68 for the non-seizure group with a p-value of 0.051. **B** Mean daily living skills domain standard scores were 63.27 for the epilepsy group and 75.36 for the non-seizure group with a p-value of 0.093. **C** Mean socialization domain standard scores were 63.18 for the epilepsy group and 69.82 for the non-seizure group with a p-value of 0.331. * p < 0.05

**Figure 8:**
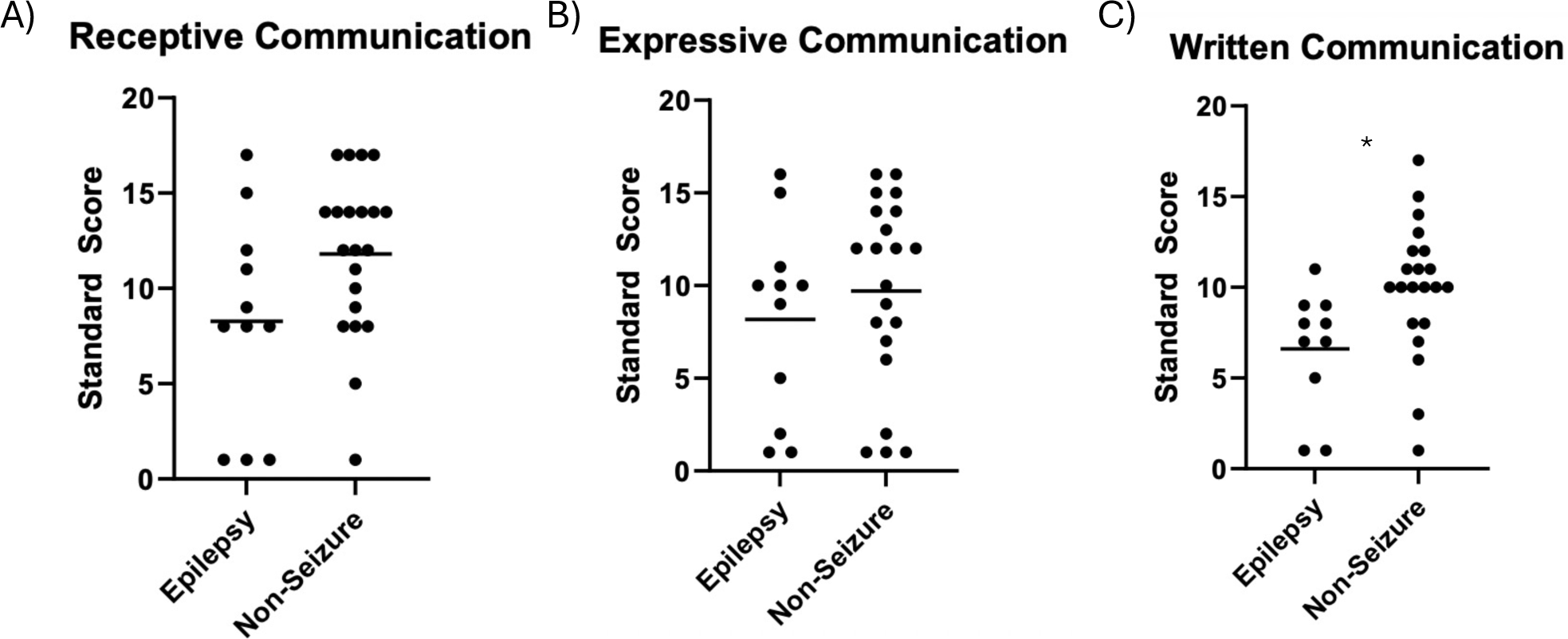
C**o**mmunication **Subdomain Scores of Epilepsy vs. Non-Seizure Groups.** The communication domain is further subdivided into receptive, expressive, and written subdomains. The normative mean for each subdomain is 20 with a standard deviation of 3. **A** Mean receptive communication subdomain standard scores were 8.27 for the epilepsy group and 11.81 for the non-seizure group with a p-value of 0.051. **B** Mean expressive communication subdomain standard scores were 8.18 for the epilepsy group and 9.71 for the non-seizure group with a p-value of 0.430. **C** Mean written communication subdomain standard scores were 6.60 for the epilepsy group and 9.95 for the non-seizure group with a p-value of 0.021. Written communication displayed statistical significance between groups, suggesting increased likelihood of those with KBG syndrome and epilepsy demonstrating worse ability to communicate through writing than those with KBG without epilepsy. * p < 0.05

**Figure 9:**
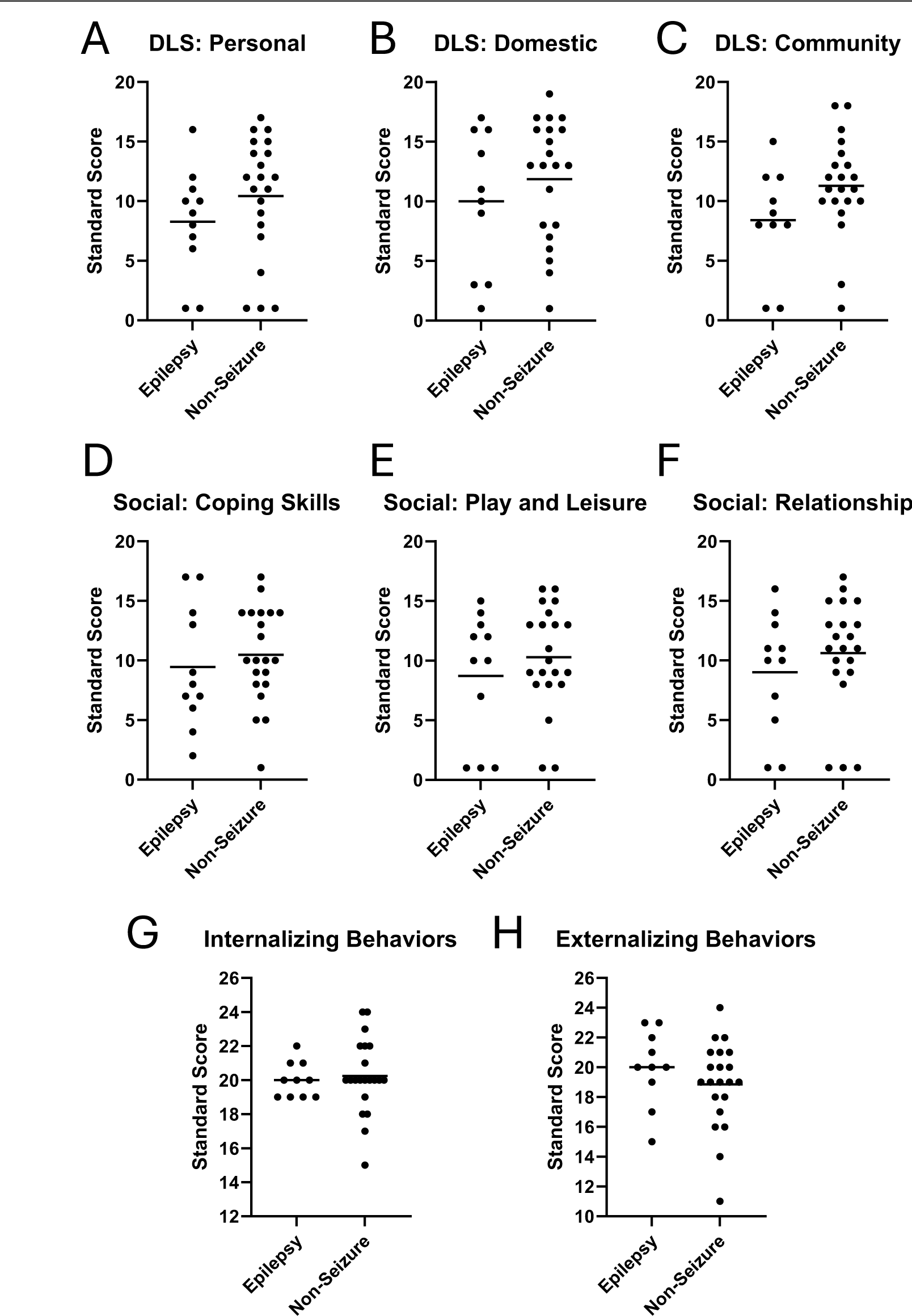
A**d**ditional **Subdomain Scores of Epilepsy vs. Non-Seizure Groups.** The daily living skills, socialization, and maladaptive behavior domains are each divided into subdomains. The normative mean for each subdomain is 20 with a standard deviation of 3. **A** Daily living skills personal subdomain epilepsy mean = 8.27, non-seizure mean = 10.43. p-value = 0.245. **B** Daily living skills domestic subdomain epilepsy mean = 10.00, non-seizure mean = 11.86. p-value = 0.375. **C** Daily living skills community skills subdomain epilepsy mean = 8.40, non-seizure mean = 11.29. p-value = 0.087. **D** Socialization coping skills subdomain epilepsy mean = 9.46, non-seizure mean = 10.48. p-value = 0.538. **E** Socialization play and leisure subdomain epilepsy mean = 8.73, non-seizure mean = 10.29. p-value = 0.382. **F** Socialization interpersonal relationships subdomain epilepsy mean = 9.00, non-seizure mean = 10.62. p-value = 0.371. **G** Maladaptive behavior internalizing subdomain epilepsy mean = 20.00, non-seizure mean = 20.24. p-value = 0.748. **H** Maladaptive behavior externalizing subdomain epilepsy mean = 20.00, non-seizure mean = 18.86. p-value = 0.298. * p < 0.05

We further analyzed five statistical outliers who had significantly lower ABC standard scores compared to the means. We discovered major shared characteristics between these individuals regarded obstetric and gynecological complications during childbirth. Of these five individuals with significantly lower Vineland scores, three have a diagnosis of epilepsy (KBG-GJL-003, KBG-GJL-34, and KBG-GJL-38) **(Table 2)**. All five required emergency cesarean sections, and three of the five were delivered prematurely.

One of the two outlier participants without epilepsy, KBG-GJL-014, was delivered several weeks early via cesarean section, resided in the neonatal intensive care unit (NICU) for four weeks, and remained hospitalized for several months due to aspiration and necrotizing enterocolitis. Her additional diagnoses include global developmental delays, migraines, hearing loss, and Crohn’s disease. KBG-GJL-023 was delivered via emergency cesarean section a few weeks early after shunts were placed for severe bilateral pleural effusions around 26-31 weeks of fetal age. This was followed by a month-long stay in the NICU with six thoracentesis procedures for pleural fluid drainage. This individual’s additional diagnoses include severe global developmental delays, autism, obsessive-compulsive disorder, and attention-deficit hyperactivity disorder.

Of the outliers with epilepsy, KBG-GJL-003 was delivered at full term via emergency cesarean section due to concerning fetal heart rate. Additional diagnoses include autism, attention-deficit hyperactivity disorder, global developmental delays, and epilepsy with tonic-clonic seizures diagnosed sometime between 0-4 years of age. KBG-GJL-034 was born via emergency cesarean section a few weeks early with a prolonged hospital stay. Additional diagnoses of this individual include tonic-clonic seizures, autism, and global developmental delays. KBG-GJL-038 was delivered via emergency cesarean section at full gestational age due to concerns about fetal heart rate. His diagnoses include global developmental delay, myoclonic epilepsy, autism, and significant speech regression. He is no longer verbal, despite knowing approximately 30 words at age 0-4 years of age. His mother states that she has observed regression which she believes was likely involving his seizures.

## Discussion

The aggregate cohort exhibited a Vineland–3 performance measuring two standard deviations below the standard 100 mean, indicative of cognitive impairment relative to those not afflicted with KBG syndrome. However, these Vineland–3 scores revealed a comparatively less pronounced degree of impairment than expected. In contrast to those with Ogden syndrome^19^, those with KBG syndrome scored relatively higher across all categories, with a subset of KBG individuals achieving scores at or above the standard mean of 100 in all domains and subdomains. The KBG syndrome group demonstrated a nearly equivalent distribution of outliers both surpassing and falling below their mean ABC and domain scores compared to Ogden syndrome as well as compared to the general population. Additionally, analyses of scores by age suggests a sustained level of adaptive functioning across the KBG syndrome cohort, devoid of discernible regression or marked enhancement at any point in time. Noteworthy differentials emerged between sexes, with males exhibiting not statistically significant but overall lower scores across ABC scores, all domains, and all subdomains.

While no statistically significant variance in ABC scores was discerned between the epilepsy versus non-seizure groups, a negative trend was evident across ABC scores and each of the principal domains. An analysis of the subdomains in communication, daily living skills, and socialization revealed that the scores for written communication were significantly higher in individuals without epilepsy (p = 0.021). This is supported by previous evidence suggesting those with KBG syndrome have especially significant impairments in communication.^24^ Although no significant difference in scores was found amongst the other domains, a negative trend was present with the non-seizure group scoring higher than the epilepsy group across all categories. Despite the consistent trend of superior performance among those without epilepsy, we theorize that the overall absence of statistical significance may imply a constraint imposed by sample size.

Given the rarity of KBG syndrome, we anticipate that the statistical power of our study is limited by the size of our cohort. Evidence supporting this theory involves a family identified as KBG- Family-24, consisting of a mother and her two sons. In the phase 1 videoconferences, this family, all devoid of seizure occurrences, was initially considered for inclusion in our cohort, but it was ultimately determined that this family harbored a variant of uncertain significance (VUS) with unclear indication as to whether their missense mutation was connected to KBG syndrome. Had these three individuals been incorporated into the cohort and categorized within the non-seizure group, statistical significance would have been observed across all domains as well as the overall ABC score. This statistical significance would suggest superior performance of the non-seizure group relative to the epilepsy group in functioning.

When considering the cohort of 39 participants, 18 required C-sections, much higher than the national average C-section rates in our participants’ countries of origin, as we already reported. ^25^ Given this finding, further research should study the possible effects of C-sections and perinatal complications on adaptive functioning, as this could potentially indicate the need for earlier neurocognitive assessment and intervention based on the methods or complications of a child’s birth.

Although epilepsy is a well-described feature of KBG syndrome, the epilepsy phenotypes in our cohort were widely variable, ranging from focal to generalized seizures and from motor to non-convulsive semiology.^26^ The wide range of seizure categories in our epilepsy group resulted in a very small samples if the epilepsy group was broken down by the types of seizures experienced. The limited number of people experiencing each of the major seizure types serves as a major limitation for our study, as different seizure types and severities may alter neurocognitive outcomes in different ways. We recommend further research to delve more into types of seizures experienced. Buijsse et al.^7^ discovered that those with simultaneous KBG and epilepsy diagnoses more often had moderate to severe intellectual disability compared to those without a history of epilepsy. Their finding is significant given that seizures occur in a large portion of the KBG syndrome population (up to 33%), with onset occurring during infancy through mid-teens and often recurring at or shortly after adolescence.^27^ Moreover, previous studies have cited high rates of remission of seizures and discontinuation of anti-epileptic medications in the KBG population up to 55% (28); however, within our cohort nine of the 11 individuals with seizures remained on medication at the time of initial videoconferencing to control their seizure disorder.

The restricted sample size remains the most relevant limitation to our study, especially displayed given the drastic change in results when three participants (KBG-Family-24) are added. This sample size weakens the power of our study and suggests that adding more participants into each group may have further stratified scores between epilepsy versus non-seizures. Additional limitations include the distribution of our cohort, as most of our participants were children and teenagers and few were adults. Sections of the Vineland are often recommended or restricted for certain ages, such as assessment of writing being typically recommended for only those over the age of three. Limitations related to the Vineland–3 administration include caregiver reporting bias and interrater variation, as respondents may not have full accuracy when describing the tasks that the KBG individual does on a daily basis. Regarding data collection, there were limited EEG files available for those diagnosed with seizures, which could provide further confirmation about the nature of seizures participants possessed.

## Conclusion

Our findings have shed light on the variation of cognitive phenotype presentations across KBG syndrome. While many individuals in our cohort experienced delays in the major Vineland domains and subdomains, others demonstrated normal functioning across these categories. When contrasted with other neurodevelopmental conditions such as Ogden syndrome, KBG syndrome exhibits an overall more favorable outcome. Furthermore, the observation that a significant proportion of individuals were delivered via C-section, including all outlier participants with the lowest Vineland scores across all categories, warrants further investigation, suggesting a potential avenue for future research.

Regarding epilepsy, our analysis underscores the significance of KBG seizure screening, as demonstrated by the observed disparities in cognitive functioning, particularly in written communication. We predict that the approaching but overall lack of statistical significance in ABC and domain scores is largely due to the low sample size of our study groups. Given these considerations, we emphasize the need for further screenings on a larger cohort, as our study is the first to utilize the Vineland–3 to assess the neurocognitive outcomes in KBG syndrome.

Performing behavioral assessments on a broader sample size of those with KBG syndrome could not only help us understand the influence of epilepsy but also provide valuable insight regarding this disorder’s cognitive trajectory to inform both research and clinical practice.

## Author contributions

G.J. Lyon devised the initial study concept and performed all phase 1 patient/family interviews.

K.P. Sarino, L. Guo, and E. Yi further designed the study, performed the Vineland—3 behavior scales, analyzed the data, and wrote the manuscript, with revision by G.J.L. All authors contributed to data collection and manuscript editing.

## Data Availability

All data produced in the present study are available upon reasonable request to the authors. Exome sequencing was done by clinical companies, so these underlying sequence data are private and not available. The genetic testing reports contain identifiable information, so cannot be shared.

## Acknowledgments

The authors would like to thank all the KBG syndrome participants and their families who participated in this study, along with the KBG Foundation for referrals.

## Ethical Approval

Both oral and written patient consent were obtained for research and publication, with approval of protocol #7659 for the Jervis Clinic by the New York State Psychiatric Institute - Columbia University Department of Psychiatry Institutional Review Board.

